# Artificial Intelligence Uncertainty Quantification in Radiotherapy Applications - A Scoping Review

**DOI:** 10.1101/2024.05.13.24307226

**Authors:** Kareem A. Wahid, Zaphanlene Y. Kaffey, David P. Farris, Laia Humbert-Vidan, Amy C. Moreno, Mathis Rasmussen, Jintao Ren, Mohamed A. Naser, Tucker J. Netherton, Stine Korreman, Guha Balakrishnan, Clifton D. Fuller, David Fuentes, Michael J. Dohopolski

## Abstract

**Background/purpose:** The use of artificial intelligence (AI) in radiotherapy (RT) is expanding rapidly. However, there exists a notable lack of clinician trust in AI models, underscoring the need for effective uncertainty quantification (UQ) methods. The purpose of this study was to scope existing literature related to UQ in RT, identify areas of improvement, and determine future directions.

**Methods:** We followed the PRISMA-ScR scoping review reporting guidelines. We utilized the population (human cancer patients), concept (utilization of AI UQ), context (radiotherapy applications) framework to structure our search and screening process. We conducted a systematic search spanning seven databases, supplemented by manual curation, up to January 2024. Our search yielded a total of 8980 articles for initial review. Manuscript screening and data extraction was performed in Covidence. Data extraction categories included general study characteristics, RT characteristics, AI characteristics, and UQ characteristics.

**Results:** We identified 56 articles published from 2015-2024. 10 domains of RT applications were represented; most studies evaluated auto-contouring (50%), followed by image-synthesis (13%), and multiple applications simultaneously (11%). 12 disease sites were represented, with head and neck cancer being the most common disease site independent of application space (32%). Imaging data was used in 91% of studies, while only 13% incorporated RT dose information. Most studies focused on failure detection as the main application of UQ (60%), with Monte Carlo dropout being the most commonly implemented UQ method (32%) followed by ensembling (16%). 55% of studies did not share code or datasets.

**Conclusion:** Our review revealed a lack of diversity in UQ for RT applications beyond auto-contouring. Moreover, there was a clear need to study additional UQ methods, such as conformal prediction. Our results may incentivize the development of guidelines for reporting and implementation of UQ in RT.

## Introduction

Artificial intelligence (AI) in healthcare has become increasingly important due to its potential to enhance diagnosis, treatment, and prognostic prediction [1]. A significant obstacle to the clinical implementation of AI that is receiving growing attention is a relative absence of model uncertainty quantification (UQ) [2]. The ability of an AI model to characterize and communicate its uncertainty, in other words, learning when to say “I don’t know” [3], could enhance clinician trust and facilitate the integration of AI into clinical practice [4–6].

Radiotherapy (RT) is a fundamental pillar of cancer treatment used in approximately 50% of all malignancies [7]. Due to the highly quantitative and structured nature of the RT clinical workflow, AI-based methodologies — namely, machine learning (ML) and deep learning (DL) — have been increasingly investigated to automate and improve a variety of tasks [8]. Advances in DL algorithms trained on increasingly larger, diverse datasets have allowed for impressive performance in a variety of RT-related applications such as image synthesis [9], registration [10], contouring [11], dose prediction [12], and outcome prediction [13–15]. However, despite the impressive performance of these models in research studies, to date there are relatively few standard AI-based tools that are routinely used in RT workflows. This hesitation could be partially attributed to insufficient clinician trust [16,17]. Enhanced UQ could bridge this trust gap, fostering greater confidence in AI applications within the RT field.

Conventionally there are two types of uncertainty: aleatoric and epistemic [18]. Aleatoric uncertainty arises from the noise inherent in the data. An example is the inherent variation in contour “ground truth” among radiation oncologists, each can be “right” but likely slightly different [19]. Epistemic uncertainty stems from incomplete information. For instance, a head and neck tumor contouring model may have limited exposure to certain rare malignancies (e.g., salivary gland cancer) and may generate poor contours with high epistemic uncertainty as these cases were underrepresented in model development. Models trained outside of medicine are often trained on datasets with >1 million samples [20]. Medical datasets, especially in RT, are considerably smaller [21], often ranging from hundreds to thousands of patient samples. Thus, epistemic uncertainty estimation would be particularly important for RT model development. Together, aleatoric and epistemic uncertainty account for the total predictive uncertainty [22]. Illustrative figures related to aleatoric and epistemic uncertainty concepts are shown in **Appendix A Figure A1**.

Within the UQ literature, there exists several methods for providing estimates of uncertainty. Contemporary methods for estimating uncertainty in ML often adopt a Bayesian perspective, treating model predictions as probability distributions rather than single point values. For instance, when predicting if a patient will develop xerostomia after radiation therapy, the model might output an 80% probability instead of simply stating “yes” or “no”. These probabilistic measures could enable safer model deployment in various clinical applications [22]. For example, UQ could be used in auto-segmentation for failure detection, flagging cases with a low probability of an accurate segmentation (i.e., high uncertainty) for additional clinical review. UQ methods such as Monte Carlo dropout and ensembles, which are suggested to be grounded in Bayesian principles [23,24], have surged in popularity in recent years [25]. However, emerging techniques, such as conformal prediction [26], are increasingly drawing on more traditional statistical methodologies.

Finally, worthy of note is that UQ has historically been closely linked to calibration, which measures the agreement between predicted probabilities and observed frequencies. Large-scale ML models — particularly DL models with numerous parameters — often show poor calibration, with output probabilities being higher than observed probabilities, subsequently leading to overconfident predictions [27]. UQ methods can help quantify and mitigate poor calibration; for example Monte Carlo dropout and ensembles often inherently improve confidence calibration [28,29]. For readers interested in more technical reviews on UQ concepts generally and specific to RT, we refer to comprehensive narrative works by Hullermeier & Waegeman et al. [18] and van den Berg & Meliadò [30], respectively.

While previous systematic and scoping reviews have covered the topics of UQ in healthcare generally [25,31] and in relation to medical imaging [32–34], these studies lacked any explicit focus on RT-related applications. Therefore, we conducted this scoping review to synthesize current trends for UQ in RT and provide an outlook for the future of this important research area for clinicians and researchers. An overview of our study is illustrated in **Appendix A Figure A2**.

## Materials and Methods

This scoping review was conducted in line with the reporting guidelines of Preferred Reporting Items for Systematic Reviews and Meta-Analyses extension for Scoping Reviews (PRISMA-ScR) [35]. The pre-registration for this scoping review was performed using the Open Science Foundation Generalized Systematic Review Registration template and can be found online (https://doi.org/10.17605/OSF.IO/E3DQG). We utilized Covidence [36] — a standardized web-based literature review collaboration software platform — to perform all initial study screening and data extraction.

### Eligibility Criteria

This scoping review was conducted to summarize the state of literature that implemented AI UQ for RT. We utilized the population, concept, context (PCC) framework to develop a focus question as recommended by the Joanna Briggs Institute Scoping Review Methodology Group [37]. Population was defined as human patients undergoing RT for cancer treatment, concept was defined as utilization of AI and UQ, and context was defined as RT applications (e.g., image acquisition and synthesis, tumor and organ at risk contouring, dose prediction, outcome prediction, etc.). Additional details on the PCC eligibility criteria and its integration into the search strategy are discussed in **Appendix B**.

### Search Strategy

A medical research librarian (D.P.F.) searched MEDLINE (Ovid), Embase (Ovid), PubMed (NLM), Cochrane Library (Wiley), and Web of Science Core Collection (Clarivate) from inception to November 17, 2023, with the search executed on November 20, 2023. A supplementary search of Web of Science Preprint Citation Index (Clarivate) and Google Scholar (Alphabet Inc.) from inception to December 12, 2023 was executed on December 13, 2023 in order to adequately query gray literature such as preprints and conference proceedings. After consultation with the research team, the librarian developed and tailored the search strategy to each database and selected controlled vocabulary (MeSH and Emtree) and natural language terms for the concepts of AI, UQ, and RT. No language, publication date, or other limiters or published search hedges were used. A total of 8974 results were retrieved from the five databases including an original set of 9 key articles supplied by the research team (MEDLINE = 1084; Embase = 1708; PubMed = 1154; Cochrane = 42; Web of Science Core Collection = 4358; Web of Science Preprint Citation Index = 428; Google Scholar = 200). The full search strategy inputs for each database is available in **Appendix C**. Notably, we incorporated 6 additional manuscripts that were not captured in the initial eligibility screening post-hoc via manual citation searching up to January 19, 2024; these manuscripts were principally added because they were formally indexed after the initial search date and were deemed relevant to ensure a more up-to-date review. Search results were uploaded to Covidence; after deduplication, 6017 unique results were identified for eligibility screening. The full PRISMA-ScR flow diagram is shown in **Appendix A Figure A3**.

### Study Selection

Initial screening to ensure studies broadly fit within our defined PCC framework was performed by 2 independent reviewers (K.A.W., Z.Y.K.) based on titles and abstracts. Disagreements were mediated by an independent third senior reviewer (M.J.D.). All disagreements were discussed in a group setting with the 3 reviewers; in cases where no consensus was reached the decision of the senior reviewer was implemented. A second full text review of these articles was performed to ensure all inclusion criteria were fully satisfied (additional details in **Appendix B**). Only full English-language preprints, conference proceedings, and standard peer-reviewed publications were included for this study; conference abstracts were excluded. Conference proceedings and preprints were deemed appropriate for inclusion due to their ubiquitous nature in computational fields [38]. Preclinical and animal studies were not included in this review. 56 articles were ultimately selected for final inclusion (**Appendix A Figure A3**). All screening was performed through the Covidence online platform.

### Data Extraction

Two reviewers extracted data from the final manuscripts (K.A.W., Z.Y.K.). All extractions were cross-checked by both reviewers and a final third reviewer (M.J.D.) when disagreements occurred. Data were initially extracted using a template generated in Covidence, focusing on four categories: general study characteristics, RT characteristics, AI characteristics, and UQ characteristics. General study characteristics included manuscript type, publication year, geographic location of the study authors, and code/data availability. RT characteristics included intended RT application space (e.g., contouring, dose planning, etc.), specific data types used (e.g., CT, MRI, etc.), and patient cancer type. AI characteristics included algorithmic approach, training/validation/testing sample sizes, and properties of the validation/testing (e.g., separate set, cross-validation, etc.). AI characteristics were adapted from existing related guidelines including TRIPOD [39] and CLAIM [40]. UQ attributes included application category, method type, evaluation metrics, self-described uncertainty type (i.e., aleatoric vs. epistemic), and use of quantitative or qualitative evaluation methods. UQ application categories and definitions were adapted from Kahl et al. [22] and Lambert et al. [34]. Additional specific considerations for each category in the data extraction process are described in detail in **Appendix B**.

### Analysis

The final extracted data were analyzed using Python v. 3.10. Descriptive statistics and visual plots were generated using the pandas, seaborn, matplotlib, numpy, geopandas, and squarify Python libraries. We also compared the overlap of extracted publications in our study and publications extracted in previous systematic and scoping reviews in similar topic domains (i.e., UQ in medical applications). To accomplish this, we compiled a comprehensive list of all publications referenced in these studies during the data extraction process along with their respective titles and digital object identifiers (DOIs). Initially, we attempted to automatically match DOIs from studies in our scoping review with those in the existing literature. If no DOI match was found, we proceeded to automatically compare titles using the difflib Python library, setting a sequence match ratio threshold of at least 0.75. All identified matches were then subsequently manually verified.

### Data and Code Availability

A CSV file containing the final studies and corresponding extracted data for this scoping review are made publicly available through Figshare (doi: 10.6084/m9.figshare.25535017). All Python code used in the analysis can be found on Github (URL: https://github.com/kwahid/RT_UQ_scoping_review/tree/main).

## Results

**Table 1** presents an overview of the extracted data from the final 56 manuscripts included in this review.

**Table 1.**
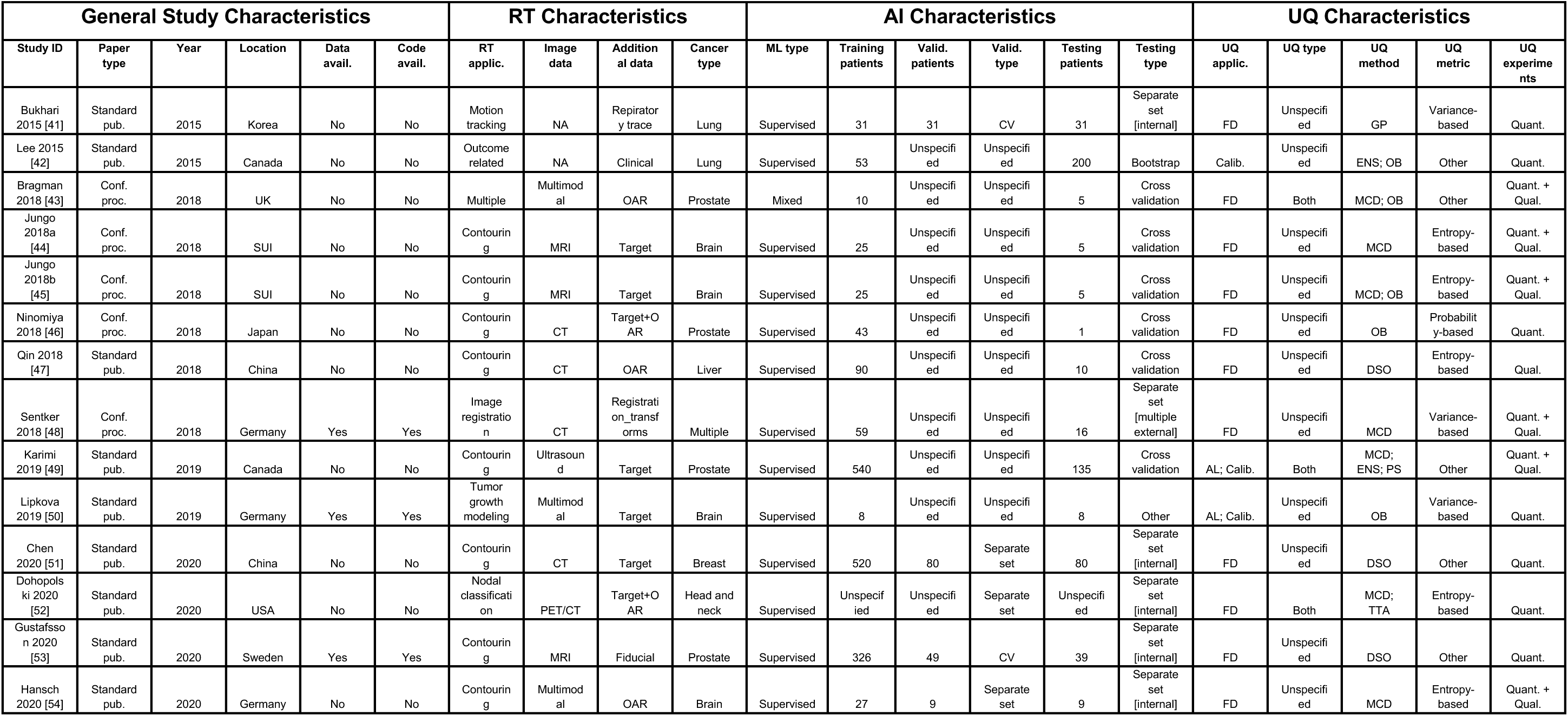
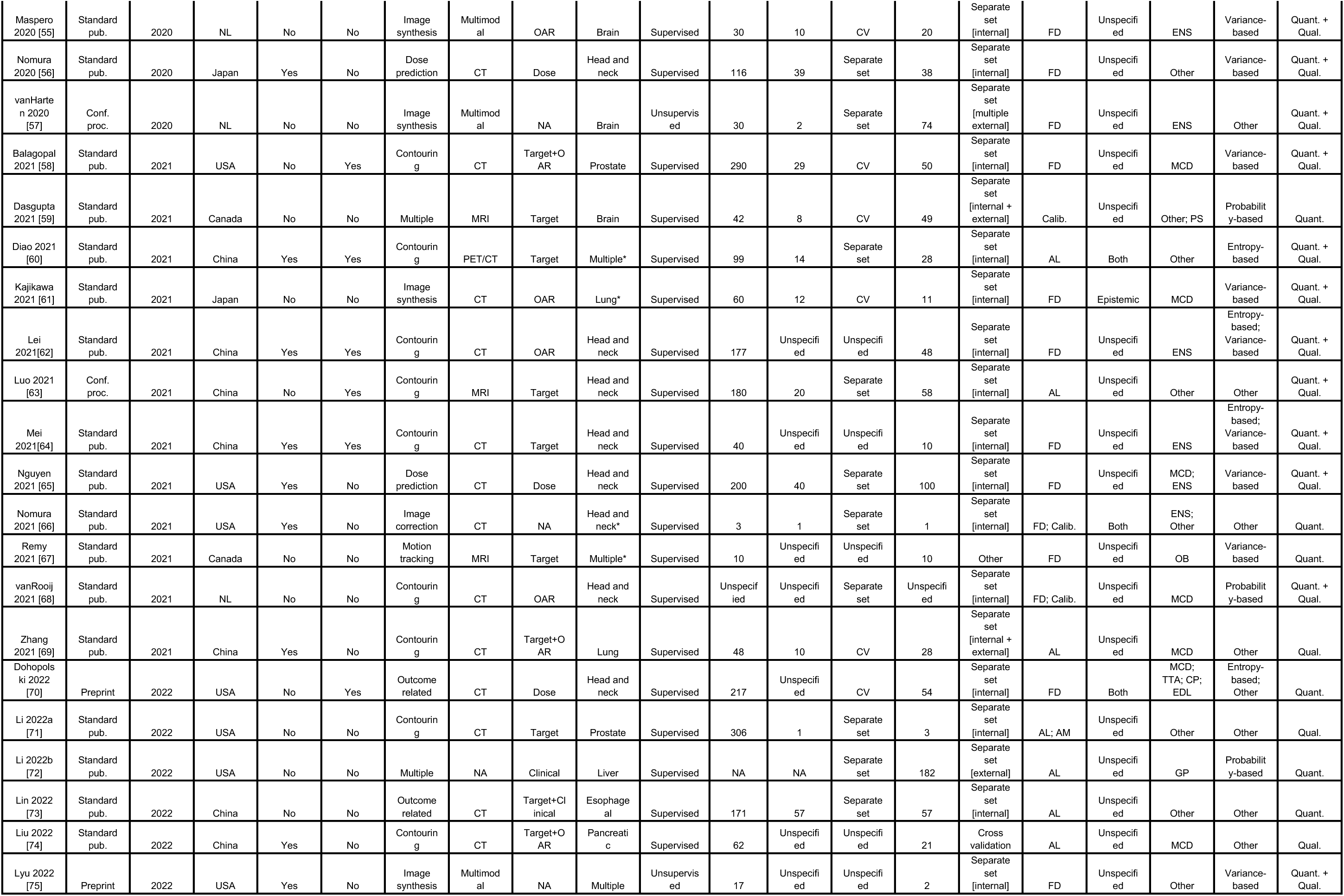
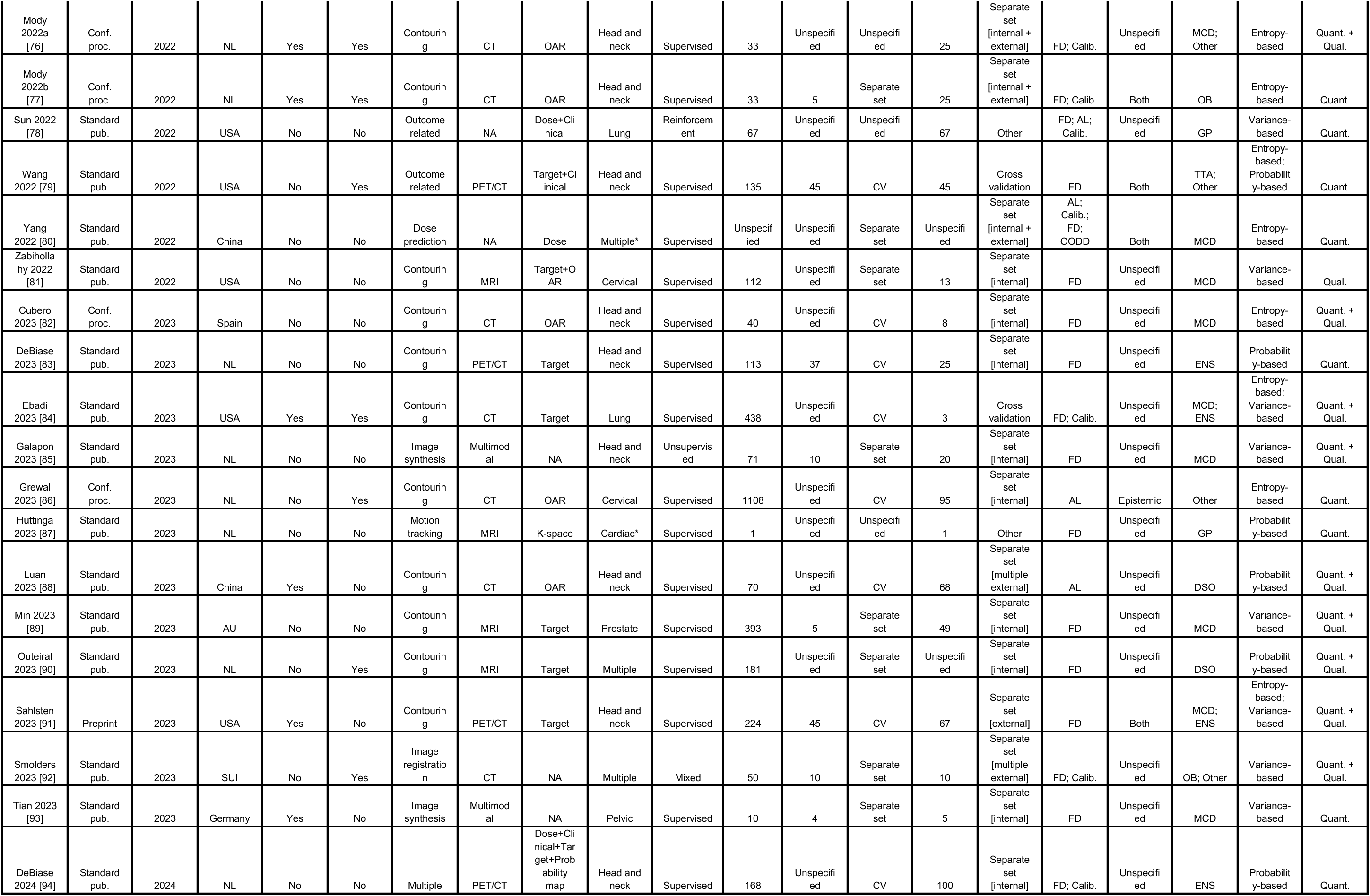
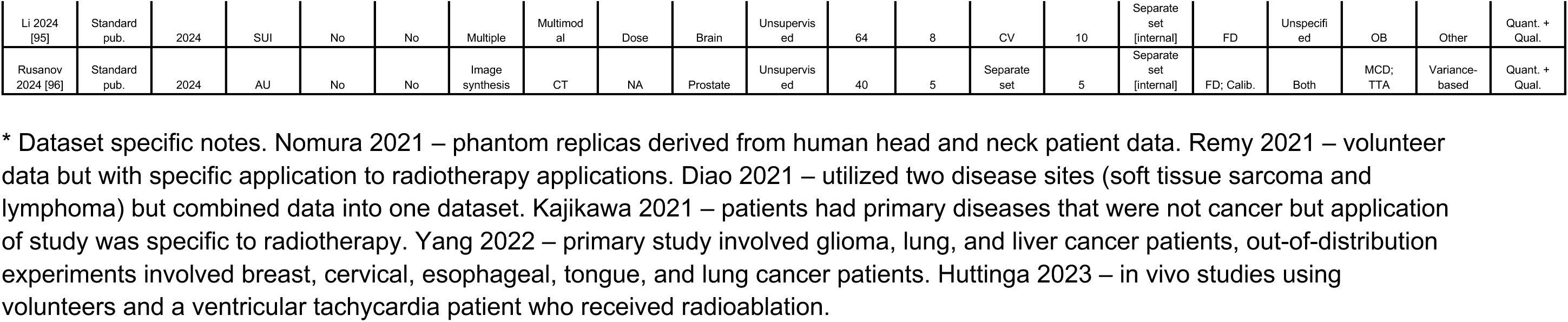
Comprehensive listing of final studies analyzed in this scoping review. Data of interest were split into four main categories: general study characteristics, radiotherapy (RT) characteristics, artificial intelligence (AI) characteristics, and uncertainty quantification (UQ) characteristics. Rows are ordered by ascending publication year and study ID. Additional abbreviations: organ at risk = OAR, machine learning = ML, cross validation = CV, failure detection = FD, active learning = AL, ambiguity modeling = AM, out of distribution detection = OODD, Gaussian Process = GP, Ensemble = ENS, Other Bayesian = OB, Platt Scaling = PS, MC Dropout = MCD, Test-time Augmentation = TTA, Evidential Deep Learning = EDL, Direct softmax output = DSO.

### General Study Characteristics

Twelve countries of origin were represented, with the majority of studies emanating from the United States (23%), China (20%), or Netherlands (20%) (**Fig 1A**, **Fig 1B**). Most studies were standard peer-reviewed publications (75%) (**Fig 1A**). The range of publication dates included in this study was 2015-2024, with most studies taking place in 2021, 2022, or 2023 (**Fig 1C**). The majority of studies did not publicly release data or code (55%), with only 32% releasing data, 29% releasing code, and 16% releasing both data and code; relative code and data availability increased in 2021, 2022, and 2023 (**Fig 1C**).

**Figure 1.**
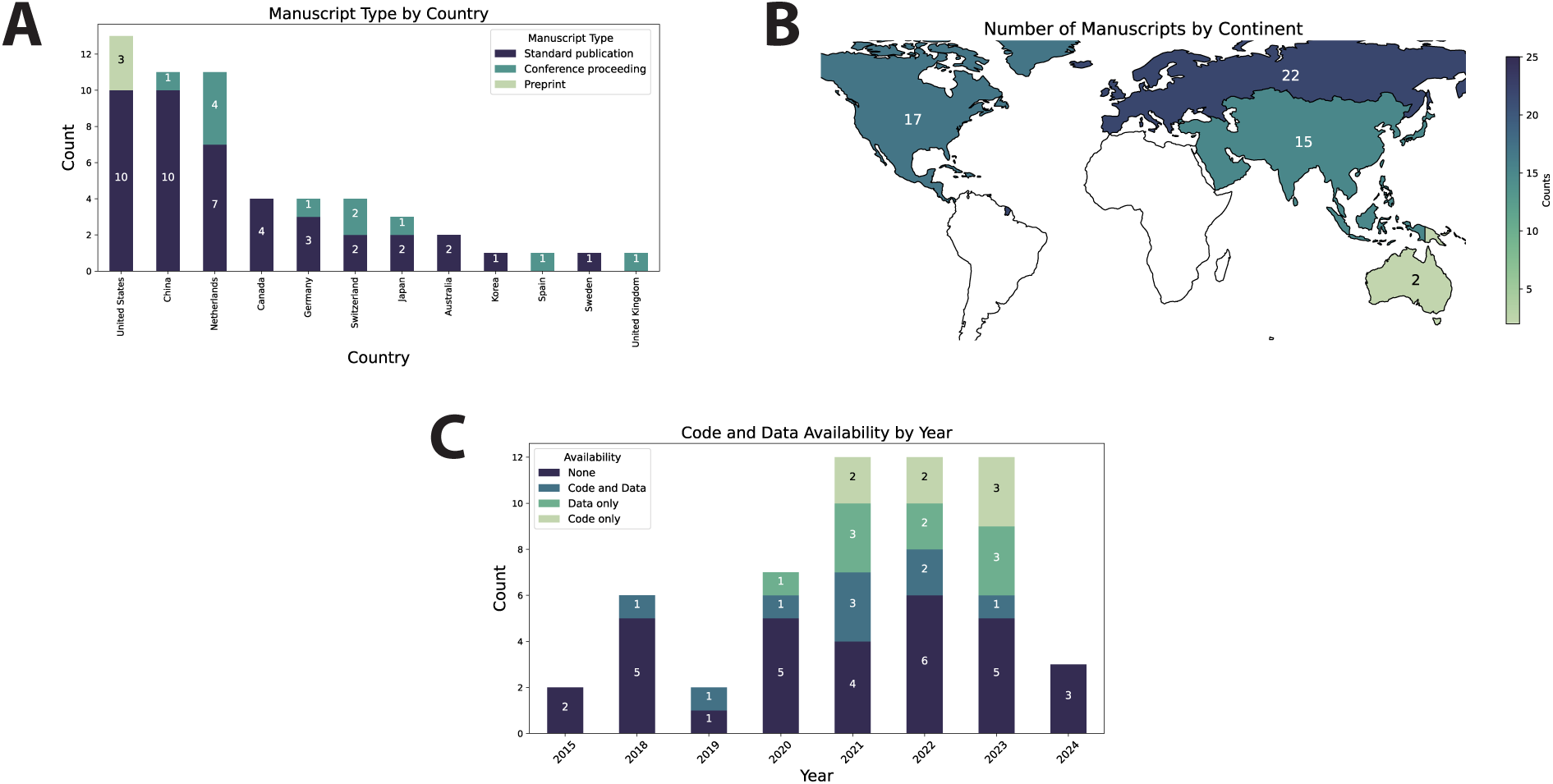
General study characteristics. (**A**) Stacked barplot showing total number of publications per country by publication type. (**B**) Heatmap of the number of studies by continent where green indicates a low number of publications and blue indicates a high number of publications; continents where no studies were extracted from are represented in white. (**C**) Stacked barplot showing code and data availability over time. Each item in the barplots corresponds to one study.

### Radiotherapy Characteristics

Multiple disease sites were included: head and neck, prostate, brain, lung, cervical, liver, esophageal, pancreatic, cardiac, breast, pelvic. Most studies were applied to head and neck cancer patient populations (32%) (**Fig 2A**). Ten RT application domains were involved: contouring, image synthesis, outcome-related, motion tracking, dose planning, image registration, nodal classification, tumor growth modeling, image correction. Most applications were focused on contouring (50%) (**Fig 2A**). Most used medical imaging data in some capacity — 45% of studies utilized CT data (**Fig 2B**); only 9% did not utilize medical imaging. The majority of studies also utilized target structures (29%), OARs (21%), or both (11%) as input data in their algorithms (**Fig 2B**); only 13% of included studies used RT dose in their algorithms.

**Figure 2.**
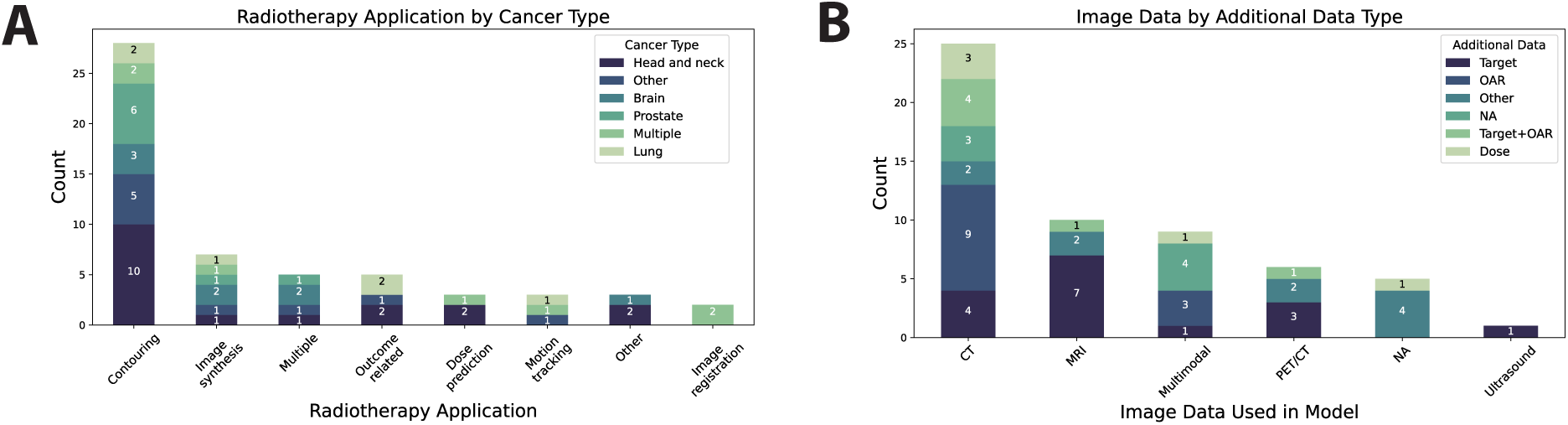
Radiotherapy characteristics. (**A**) Stacked barplot showing cancer disease site per each radiotherapy application domain. “Other” category for cancer type included cervical, liver, esophageal, pancreatic, cardiac, breast, pelvic. “Other” category for radiotherapy application included nodal classification, tumor growth modeling, and image correction. (**B**) Stacked barplot showing additional data per each imaging modality represented. “Other” category for additional data included registration transforms, respiratory trace, K-space, fiducial, clinical data, target+clinical data, dose+clinical data, and dose+clinical data+target+probability map. Each item in the barplots correspond to one study.

### AI Characteristics

The vast majority of the studies (88%) used labeled data for model training, i.e., supervised learning. Median (interquartile range) patient sample sizes were 63 (145.25), 10 (31.5), and 25 (46) for training, validation, and test datasets (**Fig 3A**). Most studies used a separate dataset for model validation (40%) compared to cross-validation approaches (30%), while 30% did not mention their validation methods. Most studies used a separate test set composed of internal, i.e., single source data (55%); only 7% of studies used multiple external validation datasets for testing (**Fig 3B**).

**Figure 3.**
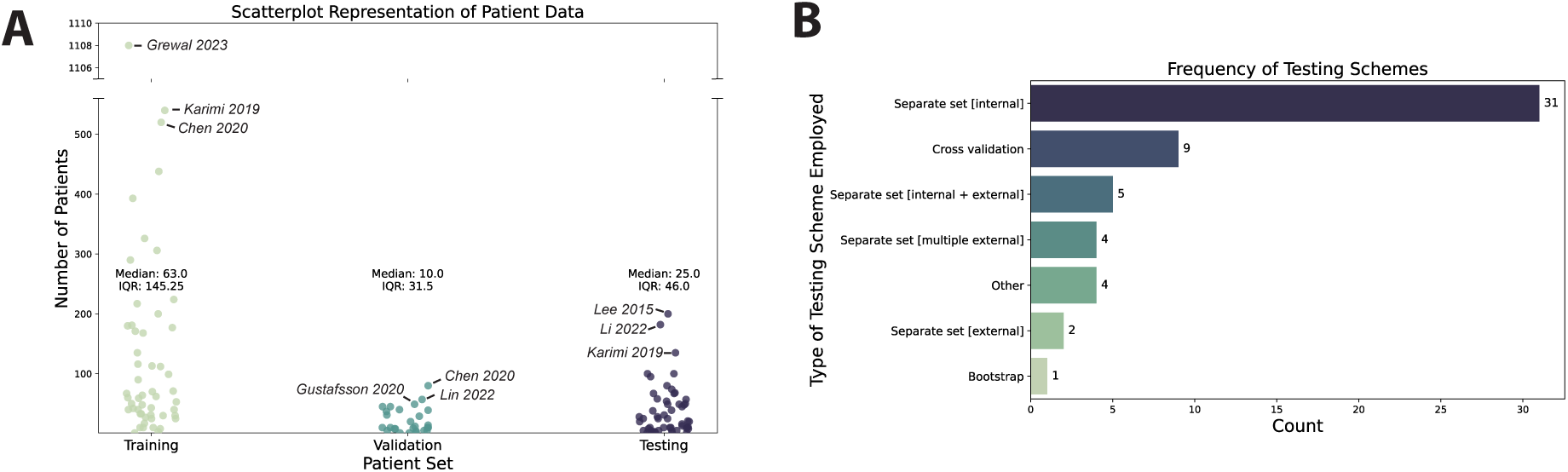
Artificial intelligence characteristics. (**A**) Scatter plot showing number of training, validation, and testing patients used in studies. Only studies that explicitly reported patient-level sample sizes are included. The three studies with the highest sample sizes in each category are annotated. (**B**) Bar plot showing types of testing strategies used in studies. Each item in the barplot corresponds to one study.

### Uncertainty Quantification Characteristics

Most studies investigated failure detection applications (60% of reported applications) followed by calibration (19%) and active learning (18%), with only a few studies investigating ambiguity modeling or out-of-distribution detection (**Fig 4A**). The majority of studies used MC Dropout (32% of reported methods), followed by ensembles (16%) and other methods (16%), with a smaller number of studies using other Bayesian methods, direct softmax outputs, test-time augmentation, gaussian processes, Platt scaling, conformal predictions, and evidential deep learning (**Fig 4B**). In terms of calculated uncertainty metrics, most studies reported using variance-based methods (34% of reported metrics) and entropy-based metrics (27%), followed by other self-defined metrics (23%), with the smallest number reporting probability based metrics (16%) (**Fig 4C**). Most studies did not explicitly report if they investigated aleatoric or epistemic uncertainty (77% of studies) and used a combination of quantitative and qualitative experiments for investigating uncertainty (52%).

**Figure 4.**
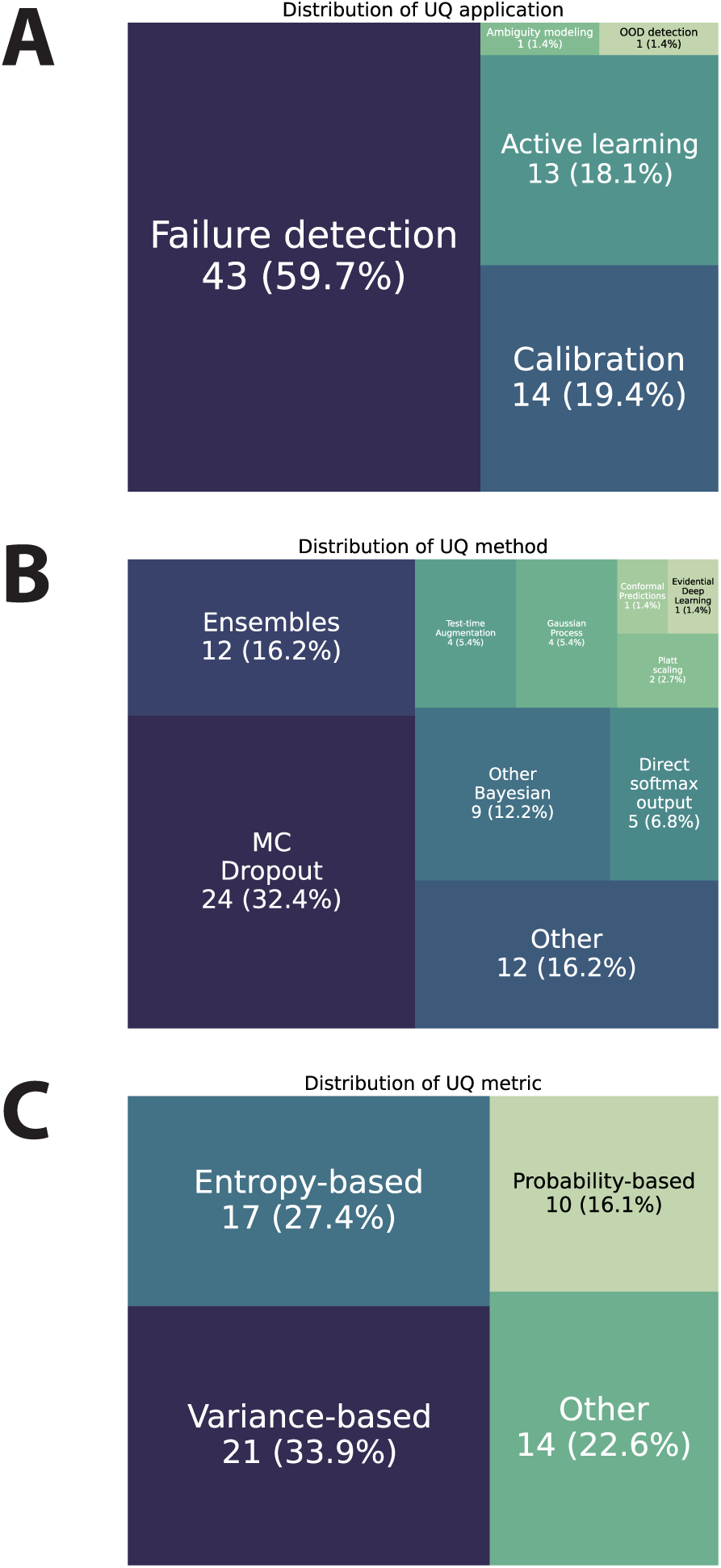
Uncertainty quantification characteristics. (**A**) Tree map of uncertainty quantification applications represented in the studies. (**B**) Tree map of uncertainty quantification methods represented in the studies. (**C**) Tree map of uncertainty quantification metrics represented in the studies. Each item in the tree maps correspond to a reported item (could be multiple per study).

### Study Overlap with Previous Reviews

Five systematic/scoping review papers related to UQ in medicine were selected for the overlap comparison. Only six studies investigated in these review papers overlapped with our 56 extracted manuscripts (**Table 2**).

**Table 2.** Study overlap with previous systematic and scoping reviews. Papers contained in previous systematic/scoping reviews related to uncertainty quantification in medicine were compared with papers extracted for our scoping review. Only final papers that were used for data extraction were compared.

| Review Paper | Overlapping Citations/Total Citations (%) | Specific Overlapping Manuscript(s) |
| --- | --- | --- |
| Zou et al. (2023) [32] | 3/56 (5%) | Jungo et al. (2018) [44], Jungo et al. (2018) [45], Bragman et al. (2018) [43] |
| Kurz et al. (2022) [33] | 0/22 (0%) | None |
| Lambert et al. (2024) [34] | 4/217 (2%) | Jungo et al. (2018) [44], Jungo et al. (2018) [45], Balagopal et al. (2021) [58], Mei et al. (2021) [64] |
| Loftus et al. (2022) [31] | 0/30 (0%) | None |
| Seoni et al. (2023) [25] | 2/144 (1%) | Jungo et al. (2018) [45], Lipkova et al. (2019) [50] |

## Discussion

The field of RT is increasingly incorporating AI into its various workflows. Although AI UQ is well-established in computer science, its adaptation to medicine and RT is still in its early stages. Incorporating UQ in AI models used in RT workflows has the potential to increase clinician confidence, helping bridge the translational gap between single institutional model development to multi-institutional clinical implementation. Our scoping review is a pioneering effort to systematically examine the application of UQ concepts within RT.

We identified several trends in UQ research in RT, likely driven by technical innovations within the broader AI research community. Mirroring practices from computer science [38], a considerable number of manuscripts were conference proceedings rather than traditional publications. Notably, we found a predominant contribution of studies from the European Union (EU). Given stringent EU data protection laws — such as The General Data Protection Regulation (GDPR) which poses challenges for secondary data use in research [97] — this raises considerations for how practitioners of UQ in RT should value data sharing. There exists a known tension between open science principles and protecting patient privacy. Although code and data availability have become standard in AI-related research, medical applications lag in this regard [98–100]. Our analysis revealed a gradual increase in code and data availability over time, reflecting a slowly evolving open science ethos in the RT community [21]. Notably, the National Institutes of Health (NIH) new Data Management and Sharing policy, effective January 2023, mandates broader data sharing for NIH-supported research [101], aligning with these open science principles. In light of these findings, we advocate for the publication of code and anonymized data in AI UQ research in RT wherever feasible to enhance reproducibility. When code or data sharing is not possible, future work should encourage privacy-preserving methodologies, like federated learning [102], as viable alternatives to conventional data sharing practices.

The extracted manuscripts covered various RT application spaces and disease sites. Auto-contouring was the most common application, aligning with its prevalence in AI-based RT [11,103,104]. Many studies focused on head and neck cancer, likely due to the complexity of this disease site, which requires precise delineation of numerous organs at risk (OARs) and challenging tumor-related target structures [105]. Most auto-contouring studies investigated OARs on CT imaging. Target structures were often generated with imaging modalities beyond CT, such as MRI for enhanced soft tissue contrast or PET for metabolic activity incorporation, matching physician practice patterns [106]. The variability observed in OARs and target structures can be characterized as aleatoric uncertainty driven by physician judgment [104,107,108]. Interestingly, Karimi et al. [49] showed that reducing aleatoric uncertainty may not be as critical as ensuring large training set sizes, at least in their specific prostate cancer target use-case. This finding suggests that, for institutional model training and fine-tuning, focusing on expanding dataset size could be more impactful than minimizing underlying contour variability (i.e., addressing factors associated with epistemic uncertainty). Of note, it has recently been suggested that RT auto-contouring performance is saturating [109], driving the need for research into additional research spaces such as UQ. Future research may benefit from exploring UQ techniques specifically tailored to address aleatoric uncertainty in auto-contouring models, considering the differences in variability between OARs and target structures.

A distinct facet in RT workflows compared to other oncologic research areas is the presence of multidimensional and complex dosimetric treatment data. DL-based dose prediction is emerging as a promising alternative to traditional knowledge-based planning approaches, offering the potential for improved accuracy, reliability, and efficiency in patient-specific plan optimization [110]. The uncertainty in DL-based dose prediction models could be critical, as it could determine when model-generated plans should be directly accepted, or if manual interventions from physicians and physicists are required to improve plan quality [80]. Surprisingly, in our review there were relatively few manuscripts directly investigating model UQ in dose prediction applications [56,65,72,80,95]. This scarcity is mirrored in outcome prediction research, where only a few studies explored dose-related toxicities [42,70,72,78], as opposed to broader oncologic outcomes like survival. Naturally, a major challenge in outcome-related research stems from the limited availability of training samples. Compared with studies that leverage granular inputs (e.g., multiple image slices representing one patient), dose-related toxicity outcomes often can only be represented at the patient level, which may explain the relative scarcity of literature.

Consistent with similar medical domains reliant on imaging [111], the majority of studies in our review employed supervised learning techniques, which involve training models on labeled data to make predictions based on new, unseen data. A minority of studies explored unsupervised learning approaches [43,57,75,85,95,96] where models learn patterns and relationships from data without explicit labels. In our review, these unsupervised methods were particularly useful for image synthesis tasks. Only one study utilized reinforcement learning [78], a technique where an agent learns to make decisions based on rewards and punishments. Regardless of the ML technique employed, training dataset sizes were generally small, with the three largest patient cohorts corresponding to auto-contouring studies (520-1108 patients) [49,51,86]. ML models often struggle with small sample sizes, especially when considering complex, multidimensional data like medical images, where models must learn intricate generalizable spatial relationships. As previously noted, this challenge is intensified when prediction outputs are restricted to broad patient-level information, such as toxicity or prognosis, with each patient representing a single data point. Notably, tasks that utilize more granular training information, like auto-contouring or image synthesis, can effectively utilize the numerous data points within each image, allowing these models to achieve reasonable performance despite the limited number of patients [109,112,113]. However, given these relatively small patient sample sizes, it is likely that intrinsic epistemic uncertainty would be high. Subsequently, carefully designed UQ may help identify patients for whom the model’s predictions are more reliable. Finally, despite the importance of using diverse and heterogeneous data for uncertainty experiments, particularly for determining how well models handle new and unknown data scenarios [22], only a handful of studies attempted to utilize multiple external test datasets [48,57,88,92]. Interestingly, this was in stark contrast to a previous scoping review on AI UQ in a broader medical context which identified a predominance of external dataset testing [31].

Ideally, UQ methods should be validated across a broad spectrum of downstream uncertainty tasks [22]. However, in our review only the study by Yang et al. [80] explored a comprehensive approach. Focusing on RT dose delivery, they evaluated several UQ applications such as active learning, calibration, failure detection, and out-of-distribution detection. Most other studies focused on singular UQ applications, with failure detection being the most common. In these studies, UQ is used as a quality assurance tool, such as flagging contours below a pre-defined quality threshold. Interestingly, model calibration, which attempts to ensure that predicted probabilities align with observed outcomes, appears somewhat underexplored in the reviewed studies, despite its historical importance in uncertainty discussions [27]. This oversight might stem from an inherent assumption that some UQ model outputs are already calibrated [28], which may not always hold true [29]. A minority of studies also used uncertainty in active learning frameworks, where the model selects the most informative data points for labeling based on their uncertainty, to improve model training. Ambiguity modeling and out-of-distribution detection were vastly underrepresented, with only one study each (Li et al. [71], and Yang et al. [80], respectively) investigating these areas.

In line with previous review literature [31,33], Monte Carlo dropout was the most frequently used UQ method in our scoping review. Monte Carlo dropout has gained widespread acceptance for its simplicity and effectiveness as a scalable approach to approximate Bayesian inference. Notably, Monte Carlo dropout, and other common methods such as ensembles often yield comparable results [114], so the superiority of specific methods is unclear and likely context-specific. In terms of uncertainty metrics, the majority of the reviewed studies favored variance-based or entropy-based metrics, aligning with their established prevalence in the literature [31,33]. There appears to be a noticeable gap in the adoption of newer, innovative approaches. For instance, only one study in our review, conducted by Dohopolski et al. on head and neck cancer outcome prediction [70], explored newer methods like evidential deep learning and statistically rigorous approaches like conformal prediction. Moreover, while qualitative analyses through heatmap visualizations were common in our extracted studies, it has been argued that conventional methods (e.g., Monte Carlo dropout, ensembles) fall short in providing spatially-correlated pixel-wise estimates [115], which may ultimately limit their clinical utility and incentivize the development of alternative approaches.

Historically, the AI UQ research community has placed significant emphasis on distinguishing between aleatoric and epistemic uncertainty. However, recent literature suggests that the ability to differentiate aleatoric from epistemic uncertainty using popular contemporary UQ techniques may not be as clear-cut as previously thought [116]. Our review revealed that the majority of studies surveyed did not explicitly identify whether their models captured aleatoric or epistemic uncertainty. This observation suggests that, at least within the RT community, the distinction between these types of uncertainty may not be deemed critical enough to warrant specific mention. Moreover, the practical significance of distinguishing between epistemic and aleatoric uncertainty may vary depending on the study’s objectives; for instance, if the primary goal is to quickly flag errors for broader human oversight (e.g., failure detection), a detailed separation of these uncertainty types might not be crucial.

Although a principal motivation behind UQ in medical AI is often believed to be the enhancement of clinician trust [5], none of the studies we reviewed explicitly investigated the influence of UQ estimates on end-user trust or decision making. This is particularly interesting given that most studies dealt with failure detection applications which would necessitate a secondary review by a clinician. Literature within diagnostic imaging applications has demonstrated that the presentation of differential outputs from an AI algorithm can impact user performance, confidence, and reliance to varying degrees [117,118]. Examining how UQ influences clinician trust and decision-making in RT through targeted human-machine interaction experiments could further elucidate the real-world impact of these tools, suggesting a vital direction for future research.

To further emphasize the novelty and need of our study, we investigated the intersection between the publications we reported on and those already cited across existing related systematic and scoping review papers. Importantly, our examination revealed just six instances of citation overlap across five distinct review papers, highlighting the originality of our research and a significant gap within the current academic discourse.

Our study, while striving for a structured and comprehensive overview of existing literature, has some limitations. Firstly, the landscape of available studies was largely limited to those indexed in the queried databases, although we supplemented our search with hand-selected literature to ensure broader coverage. Secondly, the emergent field of AI UQ presents challenges in applying traditional study quality assessment guidelines, as tailored guidelines are not yet available. However, we have taken inspiration from existing reporting guidelines, such as TRIPOD [39] and CLAIM [40], to extract relevant modeling information for our review. Furthermore, we have incorporated aspects of the newly proposed ValUES framework which aims to provide a systematic approach to validating uncertainty estimation in semantic segmentation [22], adapting its principles to enrich our review process.

Finally, an important facet of UQ that we have not considered, but should be the focus of future work is related to model bias. Notably, UQ is often part of broader discussions related to AI explainability, which is often more directly related to identifying and addressing bias. Although explainability and bias in medical AI has garnered significant attention [119], investigation of these topics in RT remains limited [17]. Moreover, while a recent small-scale study indicated that geographic biases in RT auto-contouring models are minimal [120], the necessary broader investigations across various applications have yet to be conducted. The potential for perpetuating biases and inequalities escalates when AI models function as “black boxes” with obscured decision-making processes [121]. UQ, potentially in combination with other explainability methods, could ultimately allow for improved bias detection and mitigation [6].

## Conclusions

The escalating use of UQ for RT applications signifies a key shift towards potentially more clinically impactful AI tools. Our scoping review uncovered a broad spectrum of RT applications and disease sites that have benefited from UQ. However, we observed a concentration of efforts in specific areas, such as auto-contouring, while crucial domains like dose and outcome prediction were underrepresented. Moreover, although established techniques like Monte Carlo dropout and ensembles were frequently used for failure detection applications, the exploration of alternative methods, such as conformal predictions, was limited. Notably, the majority of studies lacked code and dataset sharing suggesting a need for improved transparency and reproducibility in AI UQ research for RT. Additionally, the absence of standardized guidelines for implementing and reporting AI UQ in RT highlights a crucial area for future research. Addressing these gaps by broadening UQ applications, fostering model transparency, and developing comprehensive guidelines could significantly advance UQ in RT research.

## Data Availability

A CSV file containing the final studies and corresponding extracted data for this scoping review are made publicly available through Figshare (doi: 10.6084/m9.figshare.25535017). All Python code used in the analysis can be found on Github (URL: https://github.com/kwahid/RT_UQ_scoping_review/tree/main).
Data will be private until formal manuscript acceptance in journal.

## Acknowledgments

TJN would like to acknowledge the support of the NIH Loan Repayment Award Program.

## Declaration of generative AI and AI-assisted technologies in the writing process

During the preparation of this work, the authors used ChatGPT (GPT-4 architecture) to improve the grammatical accuracy and semantic structure of portions of the text. After using this tool, the authors reviewed and edited the content as needed and take full responsibility for the content of the publication.

## Appendices

## Appendix A: Additional Figures

**Figure A1.**
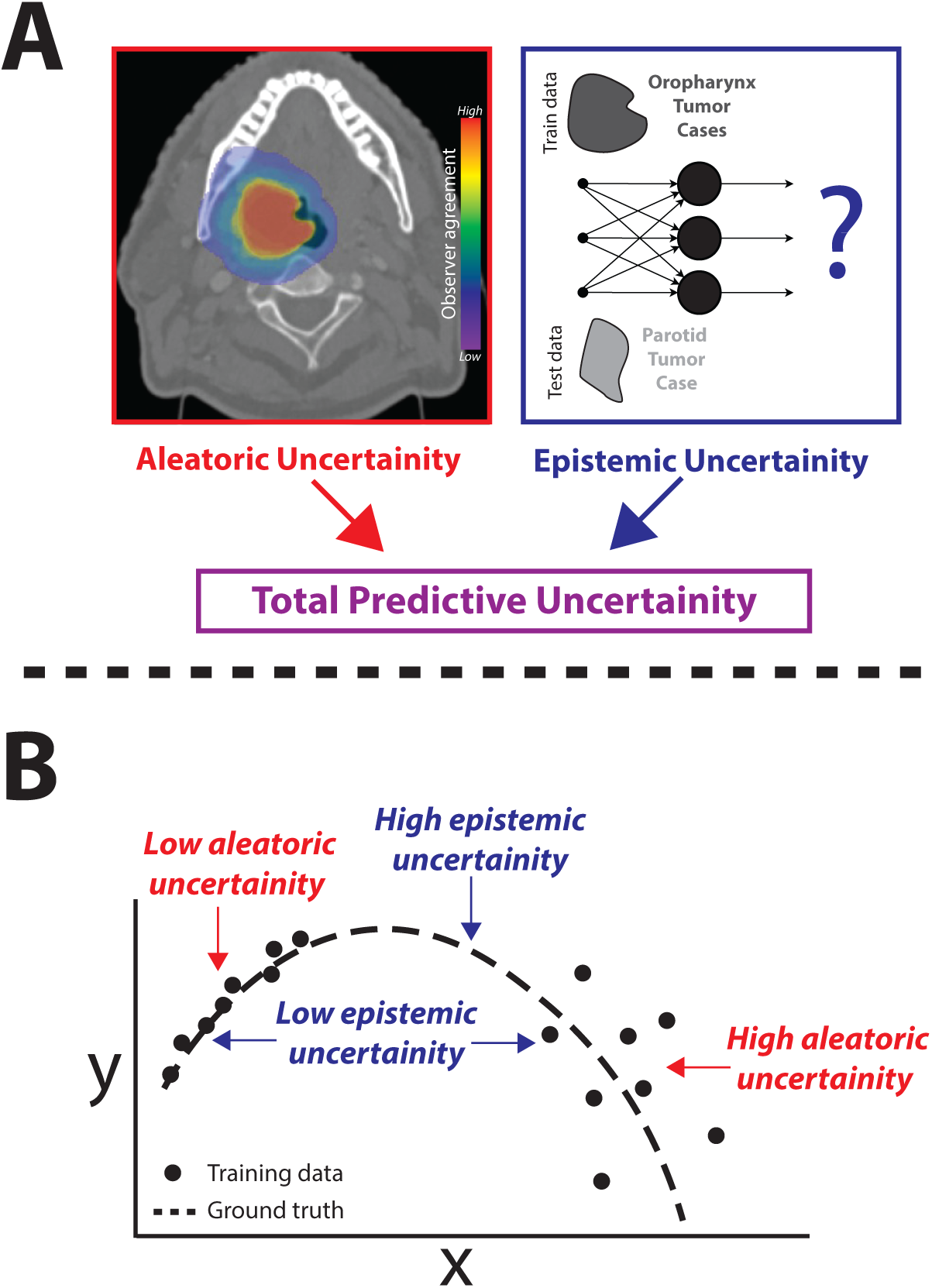
Illustrative examples of aleatoric and epistemic uncertainty concepts. (**A**) Left: A computed tomography image of an oropharyngeal cancer patient, overlaid with a probability map of interobserver agreement, illustrates aleatoric uncertainty in segmentation. Example data derived from expert contours from the Contouring Collaborative in Radiation Oncology (doi: 10.1038/s41597-023-02062-w). Right: A hypothetical tumor contouring model trained using oropharyngeal cancer cases would yield high epistemic uncertainty when presented with a parotid tumor case as a byproduct of insufficient training data. The combination of aleatoric and epistemic uncertainties contributes to the total predictive uncertainty. (**B**) A scatterplot of hypothetical variables x and y demonstrates high aleatoric uncertainty in regions with noisy data points and high epistemic uncertainty in regions with sparse data points.

**Figure A2.**
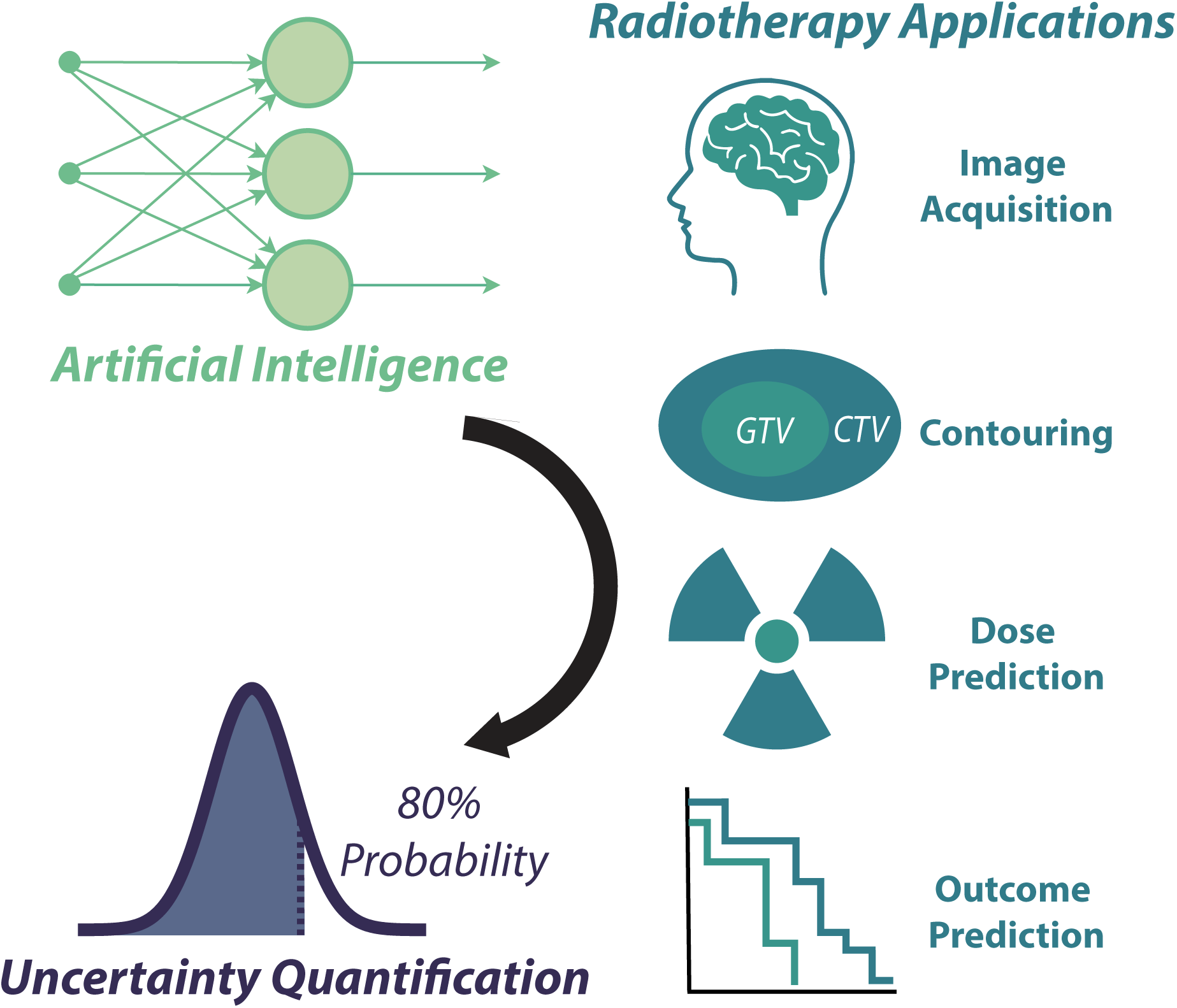
Study overview. This scoping review aims to comprehensively evaluate the literature on artificial intelligence models designed to quantify model uncertainty, specifically within the context of radiotherapy applications such as image acquisition, contouring, dose prediction, and outcome prediction, among others.

**Figure A3.**
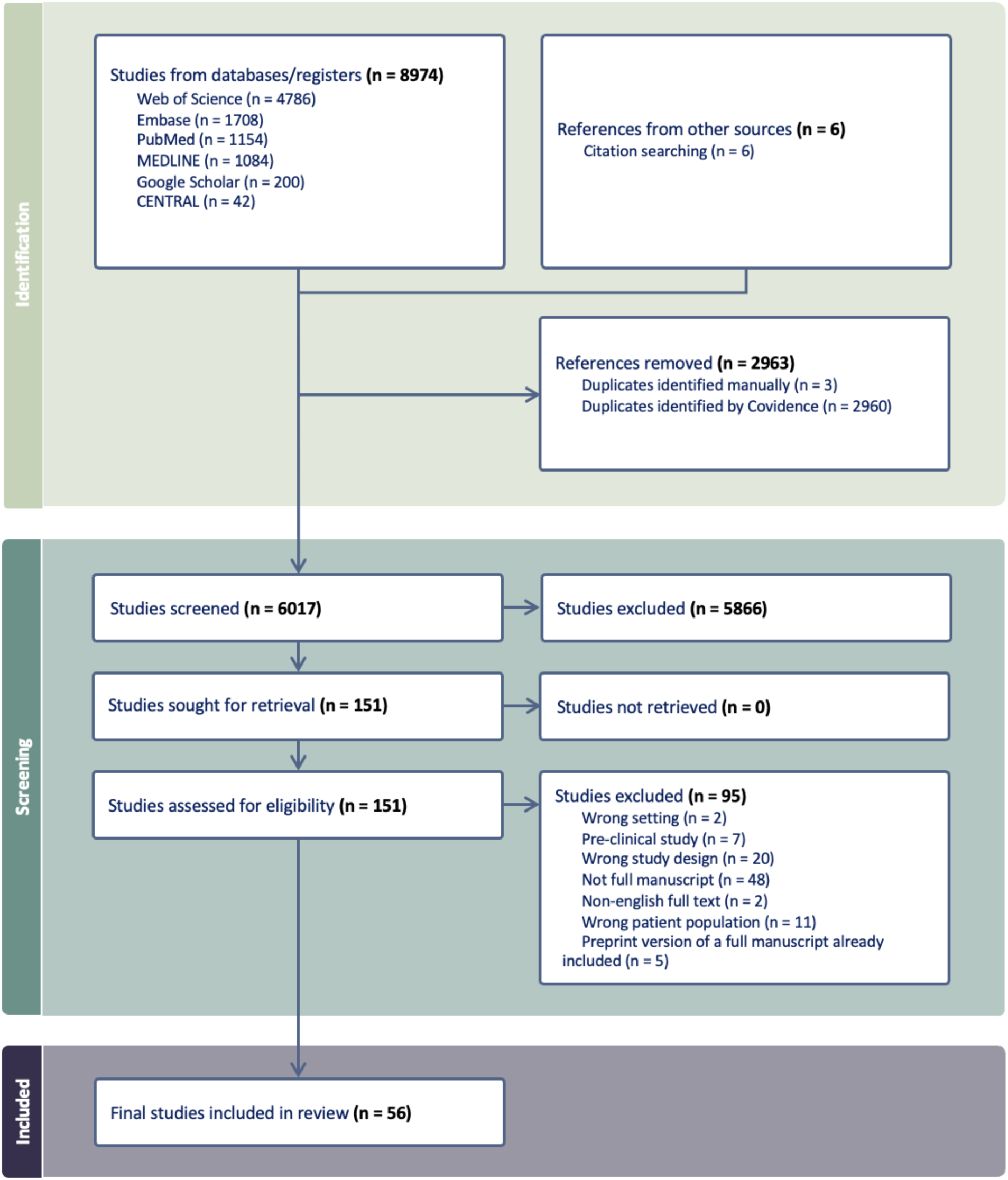
Preferred Reporting Items for Systematic Reviews and Meta-Analyses diagram illustrating systematic screening of identified studies. Ultimately, 56 studies out of the initially identified 8980 were included for the final analysis.

## Appendix B: Additional information on manuscript screening and data extraction

### Additional manuscript screening details

Two rounds of screening were performed by all reviewers which took into account all the inclusion criteria listed below. The initial screening (first round) was intended as a quick filtering process based on titles and abstracts to reduce the number of manuscripts into a workable size for eventual full-text screening (second round). Two reviewers (K.A.W. and Z.Y.K.) performed the screening process through Covidence which allows for rapid categorization of articles into inclusion/exclusion piles. Any disagreements were automatically flagged for additional review. Flagged cases underwent further scrutiny via virtual video meetings, where the two initial reviewers were joined by an independent, senior third reviewer (M.J.D.) to meticulously evaluate the contentious manuscripts. This collaborative review culminated in a final vote to decide whether the article merited inclusion. This process was repeated both for initial screening and full-text screening.

Population, concept, context (PCC) criteria for study inclusion:

1. *Population - Human patients undergoing radiotherapy for cancer treatment.* The study should explicitly mention that human patients that were actively undergoing radiotherapy, had plans to undergo radiotherapy, or had already completed radiotherapy were the subjects from which data were derived from. Studies using only preclinical samples (e.g., cell line, animal model, phantom studies, synthetic data) were excluded even if results could eventually be extrapolated to human patients. In rare circumstances where human and preclinical data was combined (e.g., mixed human data and phantom data), these studies were included. Moreover, in rare circumstances data derived from non-cancer patients were included only if directly applicable for radiotherapy specific indications (e.g., radioablation treatment of arrhythmia).
2. *Concept - Utilization of artificial intelligence and uncertainty quantification.* The study should explicitly mention that the underlying modeling technique is related to artificial intelligence or machine learning (e.g., deep learning related or more traditional methods), and must provide a method to quantify the uncertainty or confidence of the underlying model. Ideally, studies should explicitly list training and testing sample sizes, but this was not a strict requirement, particularly for older studies where this stratification was not yet standard. Studies only investigating underlying uncertainties of a radiotherapy related process (e.g., proton range uncertainty, segmentation interobserver variability, etc.) without any indication of a method to quantify predictive model uncertainty were not included.
3. *Context - Radiotherapy applications.* The study should explicitly mention that radiotherapy is the target application domain of the study or belong to a predefined list of radiotherapy applications recognized by the authors (image synthesis, image registration, contouring, dose prediction, outcome prediction). Studies in other related but distinct medical application domains (e.g., diagnostic radiology, interventional radiology, surgical oncology, medical oncology) were excluded unless the study investigated multiple applications within the same paper (e.g., diagnostic radiology applications AND radiotherapy-related applications).

Additional criteria for study inclusion:

1. Full text must be accessible to screeners. Conference abstracts must be linked to a full text (e.g., conference proceeding) or were excluded from the search.
2. Full text must be available in written English, so that it could be appropriately evaluated by all screeners.

### Additional data extraction details

Two human extractors worked in parallel to manually extract data from the final manuscripts. Specific extraction items are detailed below. These items were initially presented as a Covidence template that was used in the data extraction process and then refined if needed to fit into categorical values. All extractions were cross-checked by both reviewers (K.A.W., Z.Y.K.) and a final third reviewer (M.J.D.) when disagreements were found. Extracted data was transformed into machine readable format after initial collection based on agreement between reviewers using a version-controlled online Google Sheets document.

#### General Study Characteristics

1. Manuscript type - If the manuscript is a standard publication (i.e., published in a peer-reviewed journal), a conference proceeding (could be peer-reviewed or not), or a preprint. Articles extracted from preprint servers (e.g., arXiv) would be considered conference proceedings if explicitly indicated in the uploaded document (e.g., *this paper has been accepted to X conference*) and/or a corresponding entry was found on the conference website.
2. Publication year - Year of manuscript upload (in case of preprint) or year of publication as reported by publisher (in case of standard publication or conference proceeding).
3. Geographic location of the study authors - Which country the authors were from as determined from author affiliation information. If not all authors were from the same country, the following hierarchy was used. 1. Country where the majority of authors were from, 2. In the unlikely event of a tie, the country of the corresponding author was reported.
4. Code/data availability - If code and/or data were made publicly available. Relevant datasets DOIs and GitHub URLs were collected and reported where applicable.

#### RT Characteristics

1. Radiotherapy application space - What specific end use the manuscript is developing an artificial intelligence model for? Initially was collected with a free text option but was condensed into a categorical variable with the following possible values: dose planning, image correction, image registration, image synthesis, motion tracking, nodal classification, outcome related, contouring.
2. Specific data types used - What input data is being used for the artificial intelligence models? Initially was collected with a free text option but was condensed into two categorical variables with the following possible values:

a. Image data: CT, MRI, Multimodal, PET/CT, ultrasound, NA (i.e., none).
b. Additional data: Clinical, dose, dose+clinical, dose+clinical+target+probablity map, fiducial, K-space, organ at risk, registration transforms, respiratory trace, target, target+clinical, target + organ at risk, NA (i.e., none)
3. Cancer type of patients in the study - What were the underlying diagnoses of the patients used in the study? Initially was collected with a free text option but was condensed into a categorical variable with the following possible values: brain, breast, cardiac, cervical, esophageal, head and neck, liver, lung, multiple, pancreatic, pelvic, prostate.

#### AI Characteristics

1. Algorithmic approach - What type of underlying algorithm was used in the study? Initially was collected with a free text option but was condensed into a separate free-text variable and a categorical variable with the following possible values:

a. Machine learning type: Which overarching domain of machine learning the algorithm is categorized as: supervised, unsupervised, reinforcement, or mixed.
2. Training/validation/testing sample sizes - Specific numbers of training, validation, and testing datapoints used in the study. Data is extracted at most granular level (e.g., some algorithms use axial slices or images as input) and at the patient level. Could be NA if this information was not reported in the manuscript. We chose to focus on patient level data for reported data in our review since it was more clinically relevant and easier to compare between studies.
3. Characteristics of the validation/testing sets - How authors utilized validation and testing sets. Best practices often require separate hold-out sets but this may not always be feasible given dataset constraints. Initially was collected with a free text option but was condensed into two categorical variables with the following possible values:

a. Validation type: Cross-validation, not specified, separate set.
b. Testing type: Bootstrap, cross-validation, separate set [external], separate set [internal + external], separate set [internal], separate set [multiple external], other.

#### Uncertainty Quantification Characteristics

1. Uncertainty application category - What is the general use-case of the uncertainty methodology applied? Categories were adapted from existing literature (Kahl et al., [doi: 10.48550/arXiv.2401.08501], Lambert et al. [doi: 10.48550/arXiv.2210.03736]). Studies could investigate multiple applications simultaneously. The following specific categories were utilized:

a. Active learning: Utilization of uncertainty estimates for improving the model training process.
b. Ambiguity modeling: Comparison of model uncertainty estimates to a ground truth measure of uncertainty. For example, in segmentation, this could refer to computing the normalized cross-correlation or the generalized energy distance between the pixel-wise model measures and pixel-wise ground truth probability measures.
c. Calibration: Measurement of agreement between model estimated probabilities and true underlying data distribution probabilities. Popular methods of measuring calibration would include the Expected Calibration Error and the Brier score.
d. Failure detection: Utilize numerical model uncertainty to determine which cases should be flagged for further inspection. For example, in a segmentation framework the uncertainty estimate could be correlated to a geometric value (e.g., DSC) and subsequently binarized to classify samples below and above an expected correlated geometric value. Related to Misclassification Detection Protocol and Rejection Protocol in Lambert et al. (doi: 10.48550/arXiv.2210.03736).
e. Out-of-distribution detection: Conceptually similar to failure detection in that an uncertainty measure is used to flag cases. Typically requires a priori identification of in-distribution and out-of-distribution properties for samples (e.g., normal and abnormal images). Typically requires multiple external (out-of-distribution) datasets to implement.
2. Type of uncertainty quantification method used - Specific approach to calculate model uncertainty. Initially was collected with a free text option but was condensed into a single categorical variable. Studies could investigate multiple applications simultaneously. The following specific categories were utilized: Monte Carlo Dropout, Ensembles, Direct Softmax Output, Gaussian Process, Test-time Augmentation, Conformal Predictions, Evidential Deep Learning, Other Bayesian (an explicitly defined Bayesian approach that did not fall into a previous category), Other (bespoke approach developed in a specific paper that did not fall into a previous category).
3. Metrics used for UQ experiments - Any numerical indicators used in the computation of model uncertainty. Initially was collected with a free text option but was condensed into a single categorical variable. Studies could utilize multiple metrics simultaneously. The following specific categories were utilized: Entropy-based, Variance-based, Other (bespoke approach developed in a specific paper).
4. Self-described uncertainty type studied - Whether the study explicitly mentioned they were investigated epistemic and/or aleatoric uncertainty. Possible values of epistemic, aleatoric, both, or unspecified. Only explicit mentions of these terms (or related terms homoscedastic uncertainty and heteroscedastic uncertainty) in the manuscript were considered, otherwise this variable was labeled as unspecified (i.e., no inference about methods was performed on our part).
5. Utilization of quantitative and/or qualitative methods - Whether a uncertainty was presented in a quantitative and/or qualitative manner. Examples of qualitative experiments would include visualizing heatmap pixel-wise representations of model uncertainty in a segmentation problem.

## Appendix C: Additional information on database search criteria

### Ovid MEDLINE (R) ALL 1946 to November 17, 2023

| # | Searches | Results |
| --- | --- | --- |
| 1 | exp Artificial Intelligence/ | 183179 |
| 2 | ((artificial or machine or deep) adj (learning or intelligence)).ti,ab. | 150066 |
| 3 | ("neural net*" or "support vector" or "decision tree" or "random forest" or "gradient boost*" or bagging or ensemble or radiom*).ti,ab. | 198847 |
| 4 | or/1-3 [AI] | 370737 |
| 5 | Uncertainty/ | 18287 |
| 6 | (uncertain* or aleatoric or epistemic or "monte carlo*" or dropout or Bayes* or "conformal prediction" or "variational inference" or "temperature scaling" or platt or entropy).ti,ab. | 400538 |
| 7 | or/5-6 [Uncertainty] | 404310 |
| 8 | 4 and 7 [AI + Uncertainty] | 24799 |
| 9 | exp Radiotherapy/ | 208974 |
| 10 | exp Radiotherapy Planning, Computer-Assisted/ | 25613 |
| 11 | exp Radiation Oncology/ | 5869 |
| 12 | (radiotherap* or "radio-therap*" or irradiat* or radiat* or chemoradi* or radiochemo* or "chemo-radi*" or "radio-chemo*" or "intensity modulated" or IMRT or EBRT or photon* or proton* or radiosurgery or "radio-surgery" or brachytherapy or "brachy-therapy").ti,ab. | 1086055 |
| 13 | or/9-12 [Radiotherapy] | 1129582 |
| 14 | 8 and 13 [AI + Uncertainty + Radiotherapy] | 1084 |

### Ovid Embase Classic+Embase 1947 to 2023 November 17

| # | Searches | Results |
| --- | --- | --- |
| 1 | exp Artificial Intelligence/ | 89799 |
| 2 | ((artificial or machine or deep) adj (learning or intelligence)).ti,ab. | 176472 |
| 3 | ("neural net*" or "support vector" or "decision tree" or "random forest" or "gradient boost*" or bagging or ensemble or radiom*).ti,ab. | 234393 |
| 4 | or/1-3 [AI] | 385870 |
| 5 | Uncertainty/ | 50932 |
| 6 | (uncertain* or aleatoric or epistemic or "monte carlo*" or dropout or Bayes* or "conformal prediction" or "variational inference" or "temperature scaling" or platt or entropy).ti,ab. | 483767 |
| 7 | or/5-6 [Uncertainty] | 489765 |
| 8 | 4 and 7 [AI + Uncertainty] | 27219 |
| 9 | exp Radiotherapy/ | 736137 |
| 10 | Radiation Oncology/ | 7613 |
| 11 | (radiotherap* or "radio-therap*" or irradiat* or radiat* or chemoradi* or radiochemo* or "chemo-radi*" or "radio-chemo*" or "intensity modulated" or IMRT or EBRT or photon* or proton* or radiosurgery or "radio-surgery" or brachytherapy or "brachy-therapy").ti,ab. | 1462005 |
| 12 | or/9-11 [Radiotherapy] | 1652692 |
| 13 | 8 and 12 [AI + Uncertainty + Radiotherapy] | 1708 |

### PubMed (NLM)

((“artificial intelligence”[MeSH Terms] OR “artificial learning”[Title/Abstract] OR “artificial intelligence”[Title/Abstract] OR “machine learning”[Title/Abstract] OR “machine intelligence”[Title/Abstract] OR “deep learning”[Title/Abstract] OR “deep intelligence”[Title/Abstract] OR “neural net*”[Title/Abstract] OR “support vector”[Title/Abstract] OR “decision tree”[Title/Abstract] OR “random forest”[Title/Abstract] OR “gradient boost*”[Title/Abstract] OR “bagging”[Title/Abstract] OR “ensemble”[Title/Abstract] OR “radiom*”[Title/Abstract])

AND

(“uncertainty”[MeSH Terms] OR “uncertain*”[Title/Abstract] OR “aleatoric”[Title/Abstract] OR “epistemic”[Title/Abstract] OR “monte carlo*”[Title/Abstract] OR “dropout”[Title/Abstract] OR “bayes*”[Title/Abstract] OR “conformal prediction”[Title/Abstract] OR “variational inference”[Title/Abstract] OR “temperature scaling”[Title/Abstract] OR “platt”[Title/Abstract] OR “entropy”[Title/Abstract])

AND

(“radiotherapy”[MeSH Terms] OR “radiotherapy planning, computer assisted”[MeSH Terms] OR “radiation oncology”[MeSH Terms] OR “radiotherap*”[Title/Abstract] OR “radio therap*”[Title/Abstract] OR “irradiat*”[Title/Abstract] OR “radiat*”[Title/Abstract] OR “chemoradi*”[Title/Abstract] OR “radiochemo*”[Title/Abstract] OR “chemo radi*”[Title/Abstract] OR “radio chemo*”[Title/Abstract] OR “intensity modulated”[Title/Abstract] OR “IMRT”[Title/Abstract] OR “EBRT”[Title/Abstract] OR

“photon*”[Title/Abstract] OR “proton*”[Title/Abstract] OR “radiosurgery”[Title/Abstract] OR “radio-surgery”[Title/Abstract] OR “brachytherapy”[Title/Abstract] OR “brachy-therapy”[Title/Abstract])) Results = 1154

### Cochrane Library (Wiley)

| ID | Search | Results |
| --- | --- | --- |
| #1 | MeSH descriptor: [Artificial Intelligence] explode all trees | 2958 |
| #2 | ("artificial learning" or "artificial intelligence" or "machine learning" or "machine intelligence" or "deep learning" or "deep intelligence" or "support vector" or "decision tree" or "random forest" or bagging or ensemble or radiom*):ti,ab or ((neural NEXT net*) or (gradient NEXT boost*)):ti,ab | 7392 |
| #3 | {or #1-#2} | 8923 |
| #4 | MeSH descriptor: [Uncertainty] explode all trees | 421 |
| #5 | (uncertain* or aleatoric or epistemic or dropout or Bayes* or "conformal prediction" or "variational inference" or "temperature scaling" or platt or entropy):ti,ab or (monte NEXT carlo*):ti,ab | 28181 |
| #6 | {or #4-#5} | 28249 |
| #7 | #3 and #6 | 609 |
| #8 | MeSH descriptor: [Radiotherapy] explode all trees | 10381 |
| #9 | MeSH descriptor: [Radiotherapy Planning, Computer-Assisted] explode all trees | 467 |
| #10 | MeSH descriptor: [Radiation Oncology] explode all trees | 82 |
| #11 | (radiotherap* or irradiat* or radiat* or chemoradi* or radiochemo* or "intensity modulated" or IMRT or EBRT or photon* or proton* or radiosurgery or "radio-surgery" or brachytherapy or "brachy-therapy"):ti,ab or (((radio NEXT therap*) or (radio NEXT chemo*) or (chemo NEXT radi*))) :ti,ab | 63161 |
| #12 | {or #8-#11} | 64059 |
| #13 | #7 and #12 | 42 |

### Web of Science Core Collection (Clarivate)

Entitlements: WOS.IC: 1993 to 2023; WOS.CCR: 1985 to 2023; WOS.SCI: 1900 to 2023; WOS.AHCI: 1975 to 2023; WOS.BHCI: 2005 to 2023; WOS.BSCI: 2005 to 2023; WOS.ESCI: 2005 to 2023; WOS.ISTP: 1990 to 2023; WOS.SSCI: 1900 to 2023; WOS.ISSHP: 1990 to 2023

| ID | Search | Results |
| --- | --- | --- |
| #1 | TI=((artificial or machine or deep) NEAR/1 (learning or intelligence))<br>OR AB=((artificial or machine or deep) NEAR/1 (learning or intelligence)) | 560769 |
| #2 | TI=("neural net*" OR "neural net*" or "support vector" or "decision tree" or "random forest" or "gradient boost*" or bagging or ensemble or radiom*) OR AB=("neural net*" or "support vector" or "decision tree" or "random forest" or "gradient boost*" or bagging or ensemble or radiom*) | 988580 |
| #3 | #2 OR #1 | 1344163 |
| #4 | TI=(uncertain* or aleatoric or epistemic or "monte carlo*" or dropout or Bayes* or "conformal prediction" or "variational inference" or "temperature scaling" or platt or entropy) OR AB=(uncertain* or aleatoric or epistemic or "monte carlo*" or dropout or Bayes* or "conformal prediction" or "variational inference" or "temperature scaling" or platt or entropy) | 1533339 |
| #5 | TI=(radiotherap* or "radio-therap*" or irradiat* or radiat* or chemoradi* or radiochemo* or "chemo-radi*" or "radio-chemo*" or "intensity modulated" or IMRT or EBRT or photon* or proton* or radiosurgery or "radio-surgery" or brachytherapy or "brachy-therapy") OR AB=(radiotherap* or "radio-therap*" or irradiat* or radiat* or chemoradi* or radiochemo* or "chemo-radi*" or "radio-chemo*" or "intensity modulated" or IMRT or EBRT or photon* or proton* or radiosurgery or "radio-surgery" or brachytherapy or "brachy-therapy") | 2578649 |
| #6 | #3 AND #4 AND #5 | 4358 |

### Web of Science Preprint Citation Index (Clarivate)

| ID | Search | Results |
| --- | --- | --- |
| #1 | TI=((artificial or machine or deep) NEAR/1 (learning or intelligence))<br>OR AB=((artificial or machine or deep) NEAR/1 (learning or intelligence)) | 79524 |
| #2 | TI=("neural net*" OR "neural net*" or "support vector" or "decision tree" or "random forest" or "gradient boost*" or bagging or ensemble or radiom*) OR AB=("neural net*" or "support vector" or "decision tree" or "random forest" or "gradient boost*" or bagging or ensemble or radiom*) | 99437 |
| #3 | #2 OR #1 | 153713 |
| #4 | TI=(uncertain* or aleatoric or epistemic or "monte carlo*" or dropout or Bayes* or "conformal prediction" or "variational inference" or "temperature scaling" or platt or entropy) OR AB=(uncertain* or aleatoric or epistemic or "monte carlo*" or dropout or Bayes* or "conformal prediction" or "variational inference" or "temperature scaling" or platt or entropy) | 147982 |
| #5 | TI=(radiotherap* or "radio-therap*" or irradiat* or radiat* or chemoradi* or radiochemo* or "chemo-radi*" or "radio-chemo*" or "intensity modulated" or IMRT or EBRT or photon* or proton* or radiosurgery or "radio-surgery" or brachytherapy or "brachy-therapy") OR AB=(radiotherap* or "radio-therap*" or irradiat* or radiat* or chemoradi* or radiochemo* or "chemo-radi*" or "radio-chemo*" or "intensity modulated" or IMRT or EBRT or photon* or proton* or radiosurgery or "radio-surgery" or brachytherapy or "brachy-therapy") | 142649 |
| #6 | #3 AND #4 AND #5 | 428 |

### Google Scholar (first 200 results)

(artificial learning OR machine learning OR deep learning OR artificial intelligence OR machine intelligence OR deep intelligence OR neural network OR neural networks OR neural networking OR support vector OR support vectors OR decision tree OR decision trees OR random forest OR gradient boost OR gradient boosts OR bagging OR ensemble OR radiomic OR radiometric OR radiomorhometric) AND (uncertainty OR aleatoric OR epistemic OR “monte carlo*” OR dropout OR Bayes OR “conformal prediction” OR “variational inference” OR “temperature scaling” OR platt OR entropy) AND (radiotherapy OR irradiation OR radiation OR chemoradiation OR radiochemotherapy OR “intensity modulated” OR IMRT OR EBRT OR photon* OR proton* OR radiosurgery OR brachytherapy)

### Key Articles

To ensure a comprehensive inclusion of relevant articles, the following PubMed IDs were used as “key articles” in shaping our initial search queries: “33179605” or “33503599” or “33778184” or “34111573” or “36112996” or “36484346” or “36865296” or “37414257” or “37820691”.

